# Association of body water balance, nutritional risk, and sarcopenia with outcome in patients with acute ischemic stroke

**DOI:** 10.1101/2024.05.12.24307251

**Authors:** Takayoshi Akimoto, Kenta Tasaki, Masaki Ishihara, Makoto Hara, Hideto Nakajima

## Abstract

**Background:** Bioelectrical impedance analysis (BIA) can serve to evaluate body water balance and muscle mass. The present study aimed to explore the interrelationships among body water balance, nutritional risk, sarcopenia, and outcome after acute ischemic stroke (AIS).

**Methods:** This single-center, prospective study included 111 patients with AIS and a premorbid modified Rankin Scale score (mRS) ≤2. Overhydration, indicating a body water balance abnormality, was defined as an extracellular fluid/total body water ratio >0.390. The geriatric nutritional risk index (GNRI) was used as nutrition marker, a GNRI <92 was considered low. Sarcopenia was defined according to the 2019 Asian Working Group for Sarcopenia criteria. Poor outcome was defined as an mRS ≥3 at discharge.

**Results:** In the overall cohort, including 40 women, the median age was 77 years, 43 had poor prognosis, 31 overhydration, 25 low GNRI, and 44 sarcopenia. Patients with poor outcome had significantly higher National Institutes of Health Stroke Scale (NIHSS) scores, and significantly more common with overhydration, low GNRI, and sarcopenia (P <0.001 for all). Increasing concomitance of overhydration, low GNRI, and sarcopenia was associated with a heightened rate of poor outcomes. Especially, poor outcome was observed in 19 of 23 patients with low muscle mass and overhydration. In multivariate analysis, overhydration (odds ratio [OR] 5.504, 95% confidence interval [CI] 1.717–17.648; P = 0.004), age (OR 1.062, 95%CI 1.010–1.117; P = 0.020) and NIHSS score (OR 1.790, 95%CI 1.307–2.451; P < 0.001) were independent poor prognostic factors.

**Conclusions:** Body water balance and muscle mass assessment using BIA was useful in predicting AIS outcome. Overhydration was particularly associated with poor outcome. The combination of overhydration and low muscle mass could specifically predict poor outcomes after AIS.

**Graphical Abstract:** 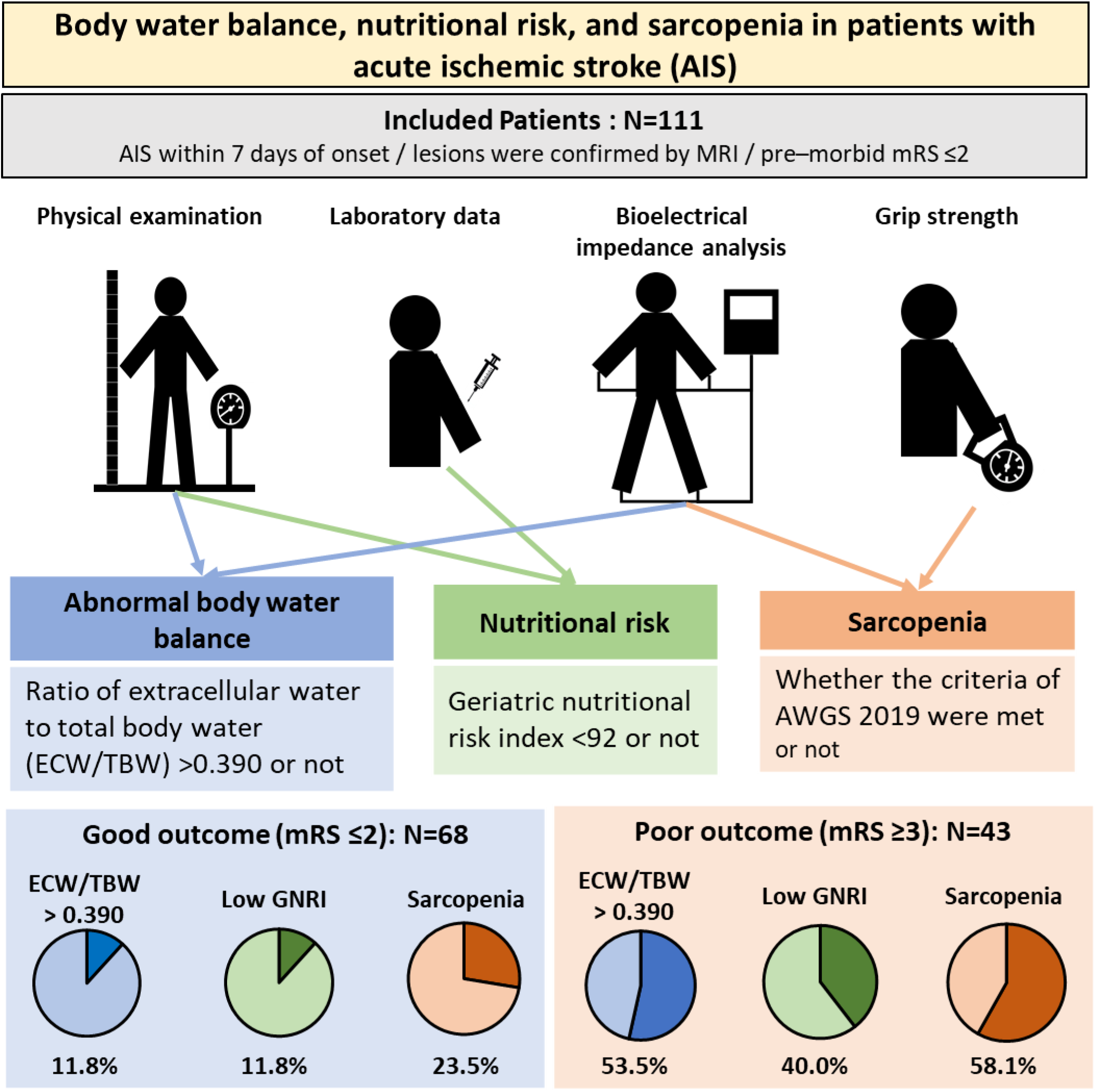

## Background

Acute ischemic stroke (AIS) is a global leading cause of death and disability.^1,2^ Given that 35% of all patients with post-stroke disability are institutionalized, improving motor function and outcomes after stroke is critical.^1^ Sarcopenia, a muscle disease that adversely impacts independence in daily living,^3,4^ has been shown to result in poor functional prognosis in patients with AIS and sarcopenia.^5–7^ According to the consensus guidelines of the European Working Group on Sarcopenia in Older People 2^4^ and the 2019 Asian Working Group for Sarcopenia,^3^ sarcopenia is defined as reduction in muscle strength or physical performance and low muscle mass or quality. Muscle strength and physical performance are evaluated using grip strength, chair-stand test, and gait speed,^3,4^ whereas skeletal muscle mass is evaluated using dual energy X-ray absorptiometry or bioelectrical impedance analysis (BIA).^3,4^ In addition to skeletal muscle mass, BIA can simultaneously measure body composition and body water balance. In human body, most water is distributed as intracellular water (ICW) and extracellular water (ECW), and total body water (TBW) is the sum of ECW and ICW. The ECW/TBW ratio indicates body water balance, and is controlled within a certain range.^8–11^ An increase in ECW/TBW indicates overhydration of body.^8–11^ Furthermore, the ECW/TBW ratio is associated with sarcopenia^12^ and undernutrition.^13,14^ Several studies in acute care hospitals have reported the relationship between the ECW/TBW ratio and stroke in the context of post-stroke dehydration,^15^ assessment of fluid replacement volume,^16^ and the stroke subtype (ischemic or hemorrhagic);^17^ however, no study to date has evaluated the association of the ECW/TBW ratio with stroke outcomes. The present study aimed to examine the usefulness of BIA for patients with AIS under acute care hospital, the interrelation among nutritionally risk, sarcopenia, and body fluid balance/overhydration, and their association with outcomes in AIS patients who were living independently before the onset of AIS.

## Methods

### Patient selection and data collection

Patients with AIS admitted to our unit between August 2021 and December 2022 and provided written informed consent were eligible for inclusion in this single-center prospective study. The AIS diagnosis was based on the presence of lesions on diffusion-weighted magnetic resonance imaging obtained within seven days of the onset of new neurologic symptoms. Patients who received alteplase or endovascular treatment were excluded. The National Institutes of Health stroke scale (NIHSS) score at admission and the modified Rankin scale (mRS) score at discharge were used to assess AIS severity. In the present study, an mRS score of ≥3 at discharge was defined as poor outcome and patients with an mRS score of ≥3 before the onset of AIS were excluded. Blood samples collected on admission were referenced. Missing values of blood collection data were not included in the analysis. All patients were evaluated for complications during hospitalization, including pneumonia, urinary tract infection, and cardiovascular complications (acute heart failure, myocardial infarction, pulmonary embolism, and deep vein thrombosis).

The study was approved by the Institutional Research Review Board of Nihon University (approval no: RK-210713-6).

### Definition of overhydration and sarcopenia

In all patients, BIA was determined in the supine resting position using Inbody S10 (InBody Co., Ltd., Seoul, Korea). ECW/TBW and muscle mass were calculated by combining BIA, height, and body weight. The estimated ECW/TBW ratio is 0.356–0.403 in healthy adults.^18–20^ The cutoff ECW/TBW ratio to define the state of overhydration varies across studies, ranging from 0.390 to 0.400.^9,13,21–23^ In the present study, to identify even mildly overhydrated patients, an ECW/TBW ratio >0.390 was used to define patients with overhydration. To avoid statistical confounding, the ECW/TBW ratio multiplied by 100 (%ECW/TBW > 39.0) was used to define overhydration, as previously reported.^24^

Sarcopenia was assessed according to the 2019 sarcopenia diagnostic criteria defined by the Asian Working Group for Sarcopenia.^3^ To prevent the effect of gait disturbance due to AIS, grip strength was used to evaluate muscle strength. Grip strength was determined using the nonparalyzed side, and decreased grip strength was defined as <28 kg for men and 18 kg for women.^3^ Low muscle mass was defined as a muscle mass of <7.0 kg/m^2^ in men and <5.7 kg/m^2^ in women.^3^ Patients with both decreased grip strength and low muscle mass were diagnosed with sarcopenia.^3^

### Definition of nutritionally at-risk

In 2018, the Global Leadership Initiative on Malnutrition criteria (GLIM) criteria were proposed as the standard diagnosis of undernutrition.^25^ Accordingly, patients considered undernourished with existing nutrition screening tools are diagnosed and classified for undernutrition severity based on weight loss rate, body mass index (BMI), and muscle mass.^25^ Since muscle mass is also used as indicator for sarcopenia in this study, the geriatric nutritional risk index (GNRI) score was employed as a nutrition-related indicator. Nutritional risk, as assessed by GNRI score, indicates a high risk of complications and death related to undernutrition.^26^ GNRI score is associated with stroke outcomes and Functional Independence Measure gains,^27,28^ and calculated by the following formula:

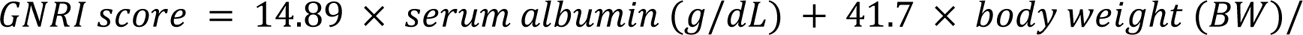

A GNRI <92 indicates nutritional risk (i.e. nutritionally at-risk).^26^ The IBW was defined as body weight at a body mass index (BMI) of 22.0 kg/m^2^ using the following formula:

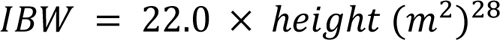

### Statistical analysis

All statistical analyses were performed using SPSS 28.0 (IBM Corp, Armonk, NY). Fisher’s exact probability test was used for nominal variables, and the Mann–Whitney *U* test was used for ordinal and continuous variables. Univariate analysis was used to categorize patients into groups according to outcome (good vs. poor outcome), overhydrated (%ECW/TBW, >39.0 vs. ≤39.0), and sarcopenia (present vs. absent). Logistic regression analysis using forward selection based on likelihood ratio tests was performed to determine the impact of overhydration, being nutritionally at-risk, and sarcopenia on the outcomes, classifying them into good and poor outcome groups. Variables with a variance inflation factor of >10 were excluded. Statistical significance was defined as a P value of <0.05.

## Results

### Study cohort

During the study period, 169 patients with AIS were admitted and 58 patients were excluded. Reasons for exclusion included disagreement with the study protocol, inadequate anthropometric measurements, and a pre-AIS mRS score of ≥3 in 41, 9, and 8 patients, respectively (Figure S1). Therefore, the final study cohort comprised 111 patients, including 40 female patients (36.0%), with a median age of 77 (19–99) years. The study dataset is available in Supplemental Materials (Table S1).

### Comparison of patients with good and poor outcomes

The characteristics of the patients categorized according to the mRS score are presented in Table 1. Briefly, 43 patients (38.7%), who experienced a poor outcome based on the study definition, were significantly older (median age, 81 vs. 73 years; P < 0.001) and had a higher NIHSS score (median score, 3 vs. 1; P < 0.001). The rate of patients with overhydration and %ECV/TBW ratio were significantly higher in the poor outcome group than in the good outcome group (53.5% vs. 11.8%, P < 0.001 and 39.2 vs. 38.2, P < 0.001, respectively). The rate of sarcopenia was 65.1% in the poor outcome group, which was significantly higher than that in the good outcome group (23.5%, P < 0.001). Furthermore, the patients in the poor outcome group had lower serum albumin (3.9 vs. 4.2 g/dL, P < 0.001) and higher N-terminal– pro-brain natriuretic peptide (NT–pro-BNP) (345.5 vs. 159 pg/mL, P < 0.001) compared to those in the good outcome group. Being nutritionally at-risk was more common in the poor outcome group than in the good outcome group (40.0% vs. 11.8%, P < 0.001). The length of hospital stay was significantly longer in the poor outcome group than in the good outcome group (21 vs. 12 days, P < 0.001).

**Table 1.**
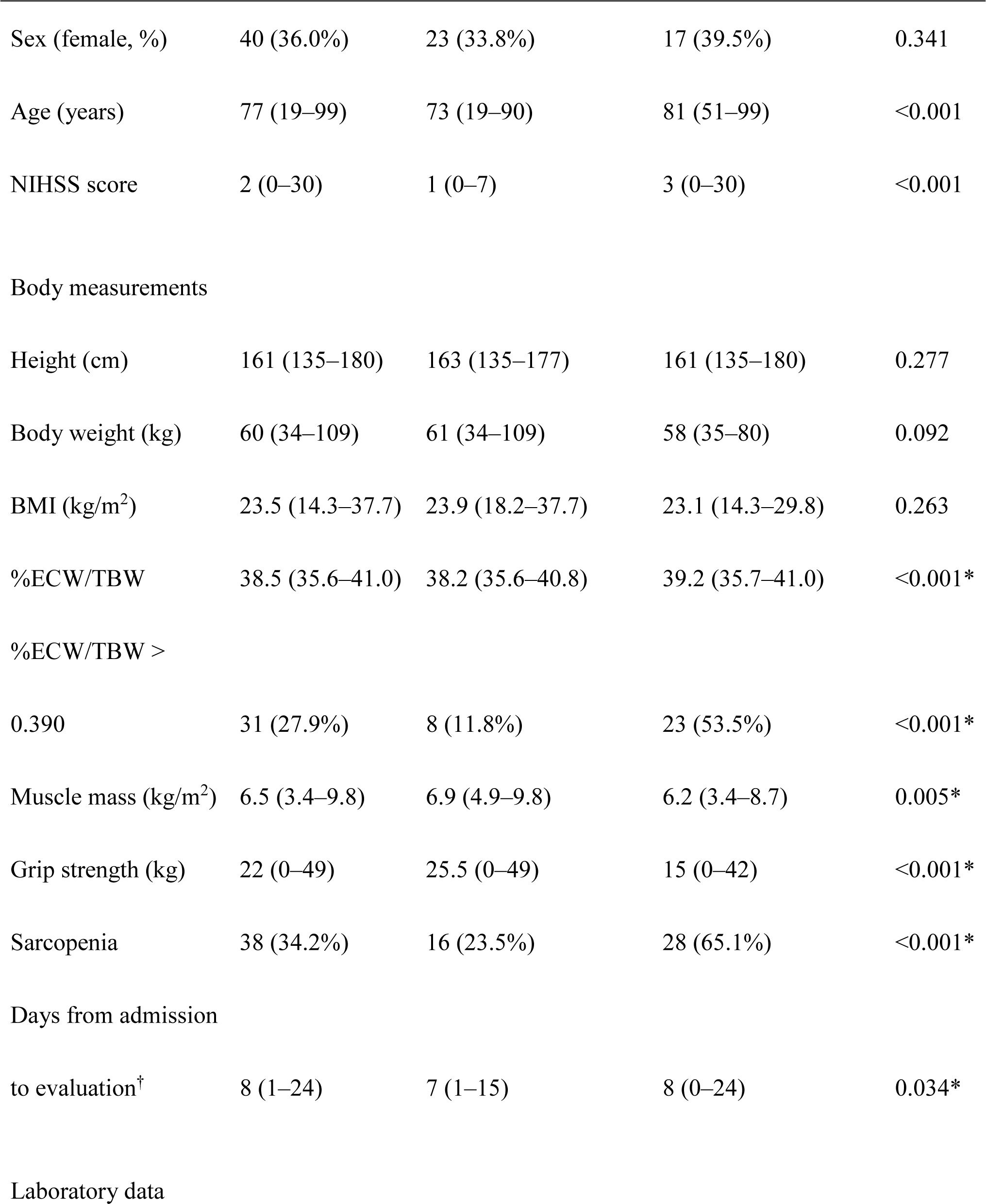
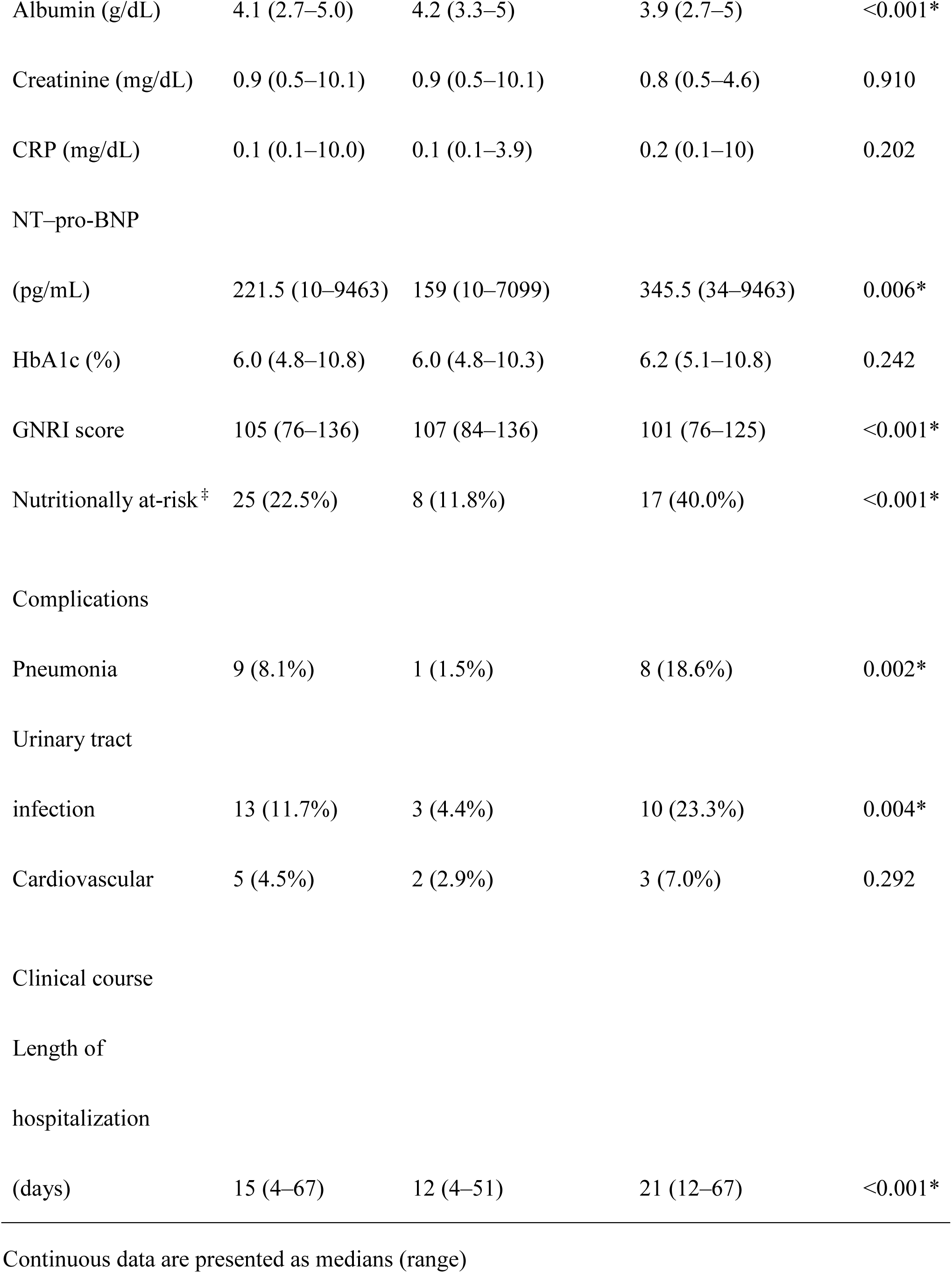
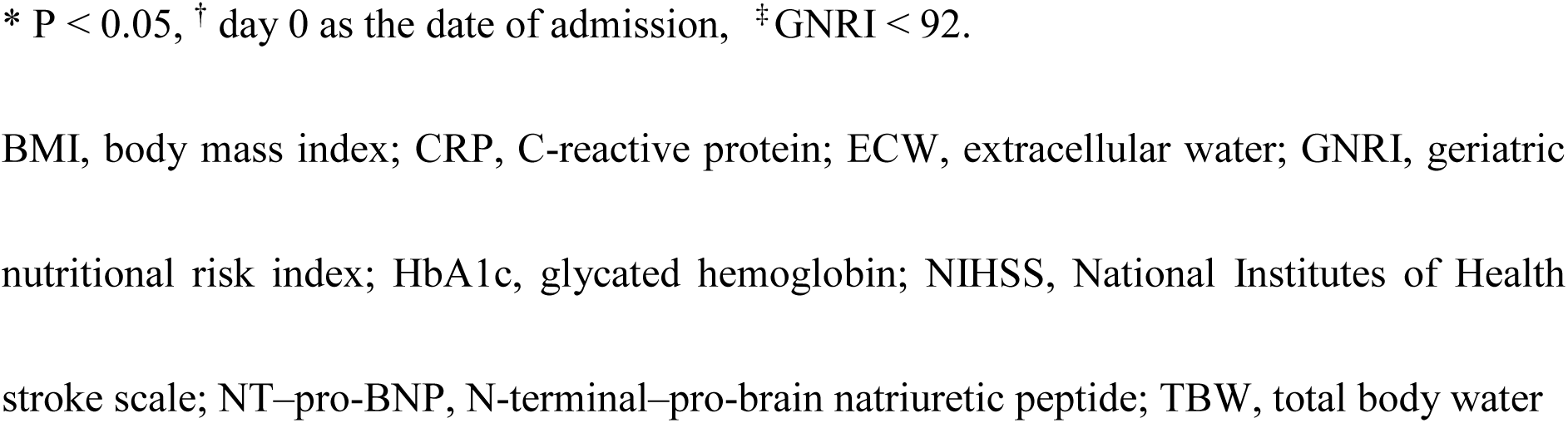
Comparison between the patients with good and poor outcomes.

|  |  | Good outcome (mRS<br>score < 3) (n = 68) | Poor outcome (mRS<br>score ≥ 3) (n = 43) | P value |
| --- | --- | --- | --- | --- |
|  | Total (N = 111) |  |  |  |
| Sex (female, %) | 40 (36.0%) | 23 (33.8%) | 17 (39.5%) | 0.341 |
| Age (years) | 77 (19–99) | 73 (19–90) | 81 (51–99) | <0.001 |
| NIHSS score | 2 (0–30) | 1 (0–7) | 3 (0–30) | <0.001 |
| Body measurements |  |  |  |  |
| Height (cm) | 161 (135–180) | 163 (135–177) | 161 (135–180) | 0.277 |
| Body weight (kg) | 60 (34–109) | 61 (34–109) | 58 (35–80) | 0.092 |
| BMI (kg/m<sup>2</sup>) | 23.5 (14.3–37.7) | 23.9 (18.2–37.7) | 23.1 (14.3–29.8) | 0.263 |
| %ECW/TBW | 38.5 (35.6–41.0) | 38.2 (35.6–40.8) | 39.2 (35.7–41.0) | <0.001* |
| %ECW/TBW > |  |  |  |  |
| 0.390 | 31 (27.9%) | 8 (11.8%) | 23 (53.5%) | <0.001* |
| Muscle mass (kg/m<sup>2</sup>) | 6.5 (3.4–9.8) | 6.9 (4.9–9.8) | 6.2 (3.4–8.7) | 0.005* |
| Grip strength (kg) | 22 (0–49) | 25.5 (0–49) | 15 (0–42) | <0.001* |
| Sarcopenia | 38 (34.2%) | 16 (23.5%) | 28 (65.1%) | <0.001* |
| Days from admission |  |  |  |  |
| to evaluation<sup>†</sup> | 8 (1–24) | 7 (1–15) | 8 (0–24) | 0.034* |
| Laboratory data |  |  |  |  |
| Albumin (g/dL) | 4.1 (2.7–5.0) | 4.2 (3.3–5) | 3.9 (2.7–5) | <0.001* |
| Creatinine (mg/dL) | 0.9 (0.5–10.1) | 0.9 (0.5–10.1) | 0.8 (0.5–4.6) | 0.910 |
| CRP (mg/dL) | 0.1 (0.1–10.0) | 0.1 (0.1–3.9) | 0.2 (0.1–10) | 0.202 |
| NT-pro-BNP |  |  |  |  |
| (pg/mL) | 221.5 (10–9463) | 159 (10–7099) | 345.5 (34–9463) | 0.006* |
| HbA1c (%) | 6.0 (4.8–10.8) | 6.0 (4.8–10.3) | 6.2 (5.1–10.8) | 0.242 |
| GNRI score | 105 (76–136) | 107 (84–136) | 101 (76–125) | <0.001* |
| Nutritionally at-risk<sup>‡</sup> | 25 (22.5%) | 8 (11.8%) | 17 (40.0%) | <0.001* |
| Complications |  |  |  |  |
| Pneumonia | 9 (8.1%) | 1 (1.5%) | 8 (18.6%) | 0.002* |
| Urinary tract |  |  |  |  |
| infection | 13 (11.7%) | 3 (4.4%) | 10 (23.3%) | 0.004* |
| Cardiovascular | 5 (4.5%) | 2 (2.9%) | 3 (7.0%) | 0.292 |
| Clinical course |  |  |  |  |
| Length of |  |  |  |  |
| hospitalization |  |  |  |  |
| (days) | 15 (4–67) | 12 (4–51) | 21 (12–67) | <0.001* |

Continuous data are presented as medians (range)
\* $P < 0.05$ , <sup>†</sup> day 0 as the date of admission, <sup>‡</sup> GNRI $< 92$ .
BMI, body mass index; CRP, C-reactive protein; ECW, extracellular water; GNRI, geriatric nutritional risk index; HbA1c, glycated hemoglobin; NIHSS, National Institutes of Health stroke scale; NT-pro-BNP, N-terminal-pro-brain natriuretic peptide; TBW, total body water

### Comparison of patients with and without overhydration

In the present study, compared with the patients without overhydration, those with overhydration (n = 31, 27.9%) included significantly more female patients (58.1% vs. 27.5%, P = 0.003) who were significantly older (83 vs. 74.5 years, P < 0.001) and had significantly higher NIHSS scores (3 vs. 1, P = 0.005) (Table 2). The rate of sarcopenia in patients with overhydration group was 74.2%, which was significantly higher than that in those without overhydration (26.3%, P<0.001). Blood tests revealed lower serum albumin (3.7 vs. 4.2 g/dL, P < 0.001) and higher NT–pro-BNP (1054 vs. 159 pg/mL, P < 0.001) levels in patients with overhydration than in those without overhydration. Being nutritionally at-risk was significantly more common in patients with overhydration than in those without overhydration (45.2% vs. 13.8%, P < 0.001). Regarding complications during hospitalization, the rate of urinary tract infection was significantly higher in patients with overhydration than in those without overhydration (22.6% vs. 7.5%, P = 0.034) whereas the rates of pneumonia and cardiovascular complications did not significantly differ between the two groups.

**Table 2.**
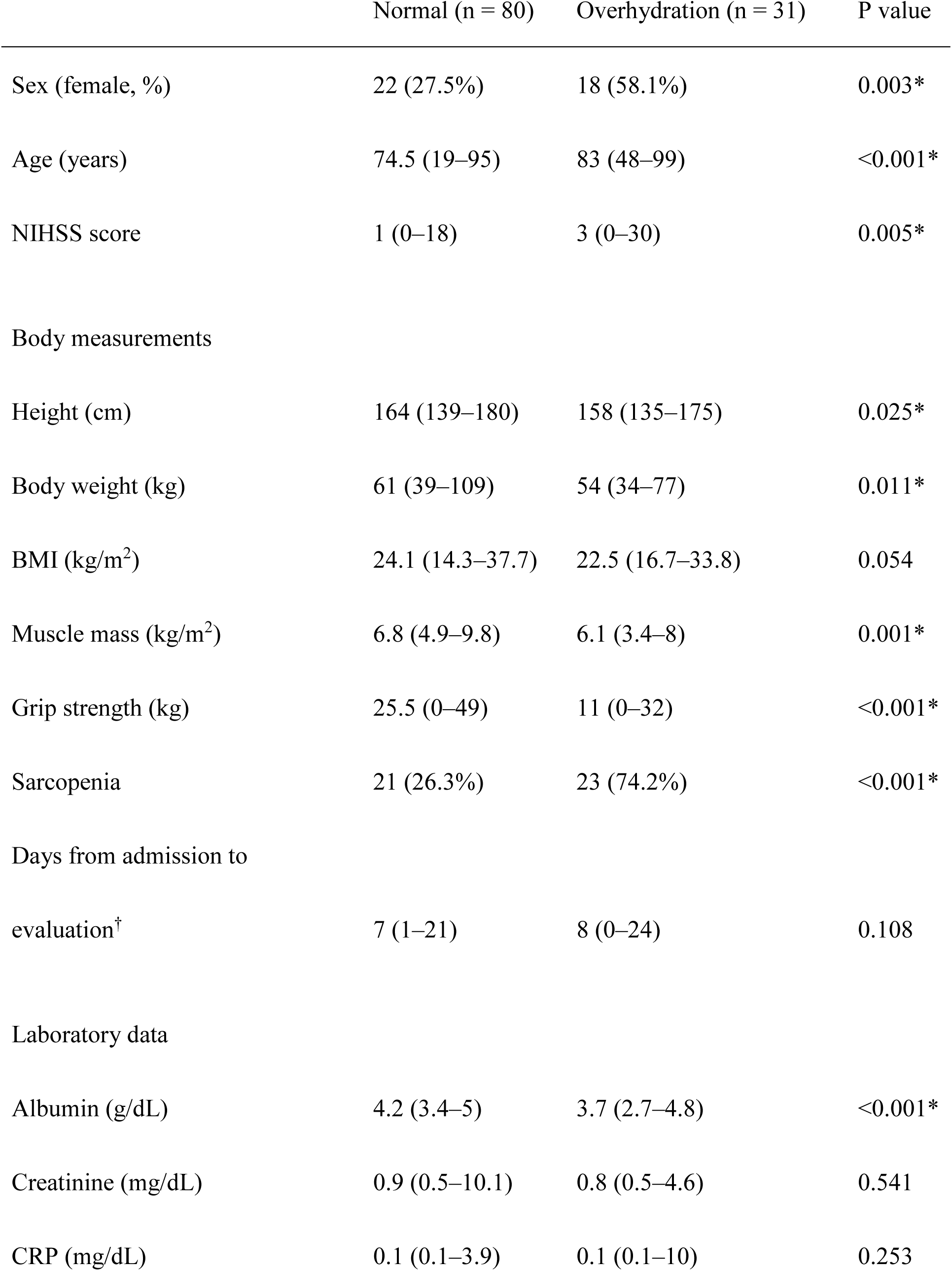
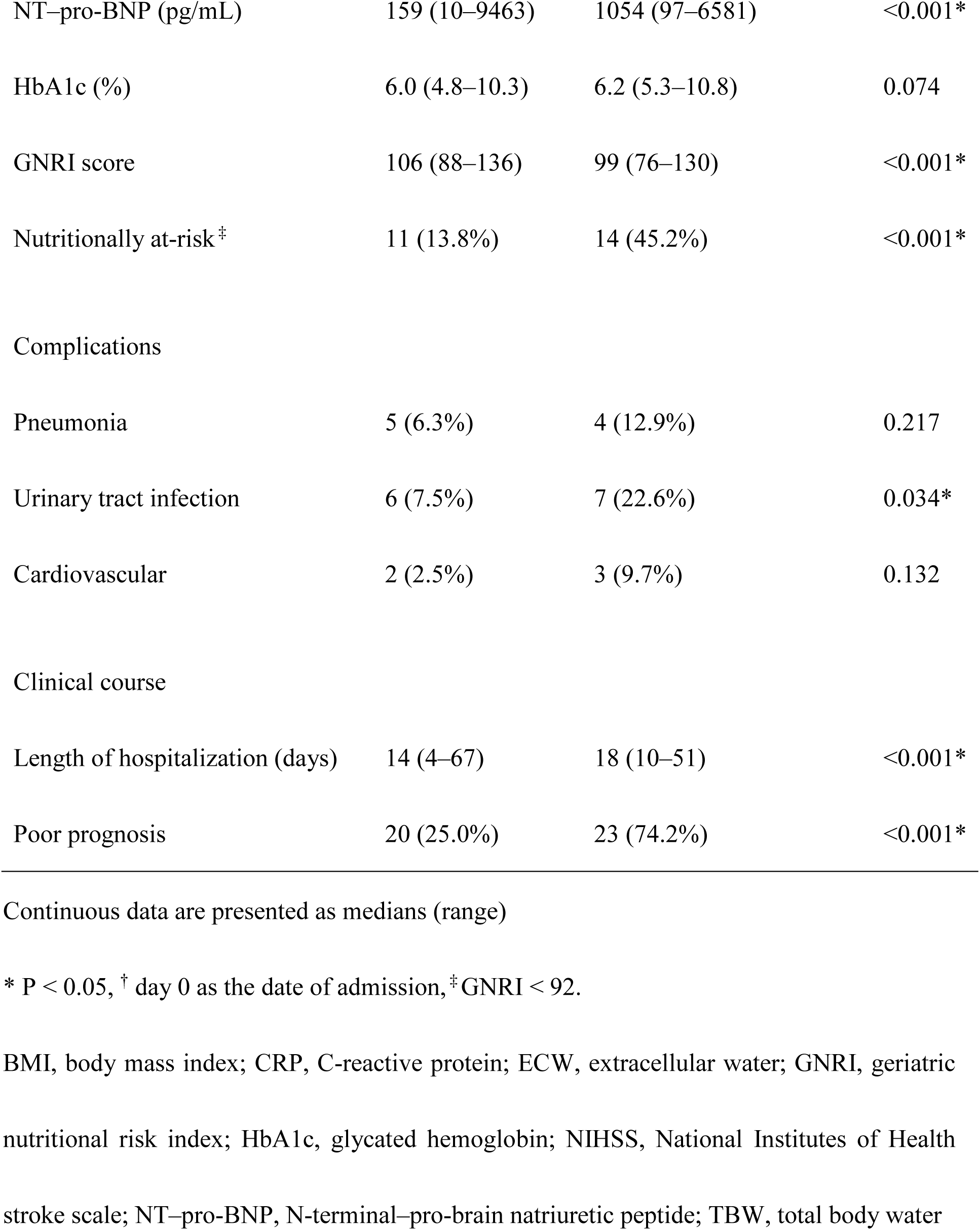
Comparison of patients with and without overhydration.

|  | Normal (n = 80) | Overhydration (n = 31) | P value |
| --- | --- | --- | --- |
| Sex (female, %) | 22 (27.5%) | 18 (58.1%) | 0.003* |
| Age (years) | 74.5 (19–95) | 83 (48–99) | <0.001* |
| NIHSS score | 1 (0–18) | 3 (0–30) | 0.005* |
| Body measurements |  |  |  |
| Height (cm) | 164 (139–180) | 158 (135–175) | 0.025* |
| Body weight (kg) | 61 (39–109) | 54 (34–77) | 0.011* |
| BMI (kg/m<sup>2</sup>) | 24.1 (14.3–37.7) | 22.5 (16.7–33.8) | 0.054 |
| Muscle mass (kg/m<sup>2</sup>) | 6.8 (4.9–9.8) | 6.1 (3.4–8) | 0.001* |
| Grip strength (kg) | 25.5 (0–49) | 11 (0–32) | <0.001* |
| Sarcopenia | 21 (26.3%) | 23 (74.2%) | <0.001* |
| Days from admission to |  |  |  |
| evaluation<sup>†</sup> | 7 (1–21) | 8 (0–24) | 0.108 |
| Laboratory data |  |  |  |
| Albumin (g/dL) | 4.2 (3.4–5) | 3.7 (2.7–4.8) | <0.001* |
| Creatinine (mg/dL) | 0.9 (0.5–10.1) | 0.8 (0.5–4.6) | 0.541 |
| CRP (mg/dL) | 0.1 (0.1–3.9) | 0.1 (0.1–10) | 0.253 |
| NT-pro-BNP (pg/mL) | 159 (10–9463) | 1054 (97–6581) | <0.001* |
| HbA1c (%) | 6.0 (4.8–10.3) | 6.2 (5.3–10.8) | 0.074 |
| GNRI score | 106 (88–136) | 99 (76–130) | <0.001* |
| Nutritionally at-risk<sup>‡</sup> | 11 (13.8%) | 14 (45.2%) | <0.001* |
| Complications |  |  |  |
| Pneumonia | 5 (6.3%) | 4 (12.9%) | 0.217 |
| Urinary tract infection | 6 (7.5%) | 7 (22.6%) | 0.034* |
| Cardiovascular | 2 (2.5%) | 3 (9.7%) | 0.132 |
| Clinical course |  |  |  |
| Length of hospitalization (days) | 14 (4–67) | 18 (10–51) | <0.001* |
| Poor prognosis | 20 (25.0%) | 23 (74.2%) | <0.001* |

Continuous data are presented as medians (range)
\* P < 0.05, † day 0 as the date of admission, ‡ GNRI < 92.
BMI, body mass index; CRP, C-reactive protein; ECW, extracellular water; GNRI, geriatric
nutritional risk index; HbA1c, glycated hemoglobin; NIHSS, National Institutes of Health
stroke scale; NT-pro-BNP, N-terminal-pro-brain natriuretic peptide; TBW, total body water

### Comparison of patients with and without sarcopenia

The comparison of patients with and without sarcopenia is summarized in Table 3. Briefly, the patients with sarcopenia (n = 44, 39.6%) were significantly older (81 vs. 72 years, P < 0.001) and had higher NIHSS scores (3 vs. 1, P = 0.001), lower BMI (21.7 vs. 24.2 kg/m^2^, P < 0.001), and higher %ECW/TBW ratios (39.2 vs. 38.1, P < 0.001). The NT–pro-BNP levels were significantly higher and the GNRI scores were significantly lower in the sarcopenia group than in the non-sarcopenia group (406 vs. 159 pg/mL, P = 0.001 and 99 vs. 107, P < 0.001, respectively). Additionally, the rate of nutritionally at-risk and pneumonia during hospitalization was significantly higher in the sarcopenia group than in the non-sarcopenia group (47.7% vs. 6.0%, P<0.001, 18.4% vs. 2.7%, P = 0.019, respectively).

**Table 3.** Comparison of patients with and without sarcopenia.

|  | Non-sarcopenia (n = 67) | Sarcopenia (n = 44) | P value |
| --- | --- | --- | --- |
| Sex (female, %) | 24 (35.8%) | 16 (36.4%) | 0.556 |
| Age (years) | 72 (19–95) | 81 (31–99) | <0.001* |
| NIHSS score | 1 (0–18) | 3 (0–30) | 0.001* |
| Body measurements |  |  |  |
| Height (cm) | 164 (139–180) | 159 (135–175) | 0.028* |
| Body weight (kg) | 64 (43–109) | 53 (34–72) | <0.001* |
| BMI (kg/m<sup>2</sup>) | 24.2 (19.0–37.7) | 21.7 (14.3–26.6) | <0.001* |
| %ECW/TBW | 38.1 (35.6–41.0) | 39.2 (36.6–40.8) | <0.001* |
| %ECW/TBW > 0.390 | 8 (11.9%) | 23 (52.3%) | <0.001* |
| Muscle mass (kg/m<sup>2</sup>) | 7.1 (5.1–9.8) | 5.9 (3.4–6.9) | <0.001* |
| Grip strength (kg) | 29 (0–49) | 16 (0–27) | <0.001* |
| Days from admission to<br>evaluation<sup>†</sup> | 7 (1–21) | 8 (0–24) | 0.058 |
| Laboratory data |  |  |  |
| Albumin (g/dL) | 4.2 (3.4–5) | 3.9 (2.7–5) | <0.001* |
| Creatinine (mg/dL) | 0.9 (0.5–10.1) | 0.9 (0.5–4.6) | 0.787 |
| CRP (mg/dL) | 0.1 (0.1–3) | 0.2 (0.1–10) | 0.080 |
| NT-pro-BNP (pg/mL) | 159 (10–7099) | 406 (34–9463) | 0.001* |
| HbA1c (%) | 5.9 (4.8–10.3) | 6.2 (5.3–10.8) | 0.323 |
| GNRI score | 107 (93–136) | 99 (76–125) | <0.001* |
| Nutritionally at-risk<sup>‡</sup> | 4 (6.0%) | 21 (47.7%) | <0.001* |
| Complications |  |  |  |
| Pneumonia | 2 (3%) | 7 (15.9%) | 0.019* |
| Urinary tract infection | 5 (7.5%) | 8 (18.2%) | 0.080 |
| Cardiovascular | 4 (6%) | 1 (2.3%) | 0.338 |
| Clinical course |  |  |  |
| Length of hospitalization |  |  |  |
| (days) | 13 (6–51) | 18 (4–67) | 0.001* |
| Poor prognosis | 15 (22.4%) | 28 (63.6%) | <0.001* |

\* P < 0.05, † day 0 as the date of admission, ‡ GNRI < 92.
BMI, body mass index; CRP, C-reactive protein; ECW, extracellular water; GNRI, geriatric nutritional risk index; NIHSS, National Institutes of Health stroke scale; NT-pro-BNP, N-terminal-pro-brain natriuretic peptide; TBW, total body water

### Relationship between body water balance and muscle mass

Figure 1 shows the scatter plots examining the relationship between %ECW/TBW and muscle mass. The study patients were categorized into the following four groups based on the ECW/TBW ratio and muscle mass using a %ECW/TBW cutoff value of 39.0^25,26^ and muscle mass cutoff values of 7.0 kg/m^2^ in men and 5.7 kg/m^2^ in women:^3^ Group A, low %ECW/TBW and high muscle mass; Group B, high %ECW/TBW and high muscle mass; Group C, low %ECW/TBW and low muscle mass; and Group D, high %ECW/TBW and low muscle mass. The overall cohort included 71 male and 40 female patients. Groups A, B, C, and D included 34 (30.6%), 1 (0.9%), 24 (21.6%), and 12 (10.8%) male patients, respectively, and 14 (35%), 7 (17.5%), 8 (20%), and 11 (27.5%) female patients, respectively.

**Figure 1.**
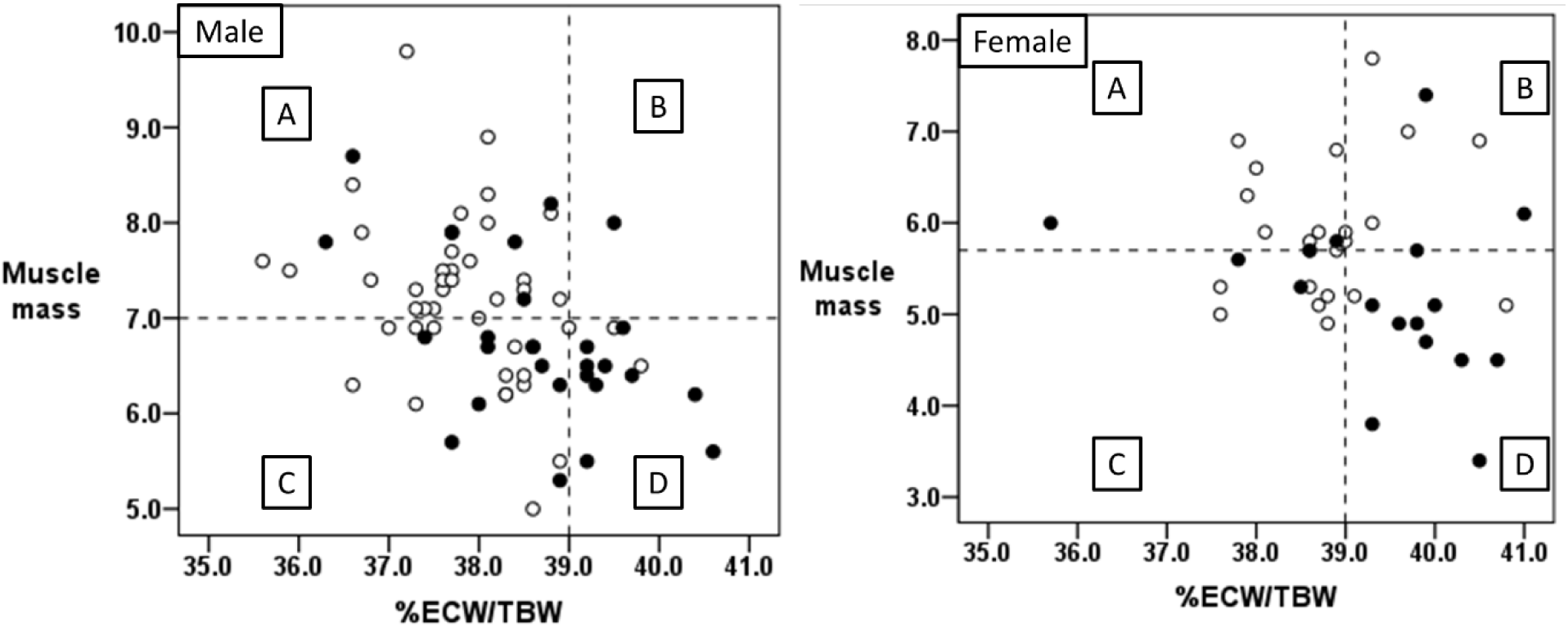
Scatter plots showing the distribution of groups categorized according to %ECW/TBW and muscle mass %ECW/TBW, ratio of extracellular fluid to total body water multiplied by 100 White and black circles indicate patients with good and poor outcomes, respectively.

In addition, we analyzed the association between outcomes and the four groups. In groups A, there were 39 patients with good outcome and nine with poor outcome, there were four of each in group B, 21 with good and 11 with poor outcomes in group C, and 4 and 19 in group D, respectively. As shown in Supplemental Table 2, the contingency table for Group D or not and outcome revealed that a high %ECW/TBW with a low muscle mass (Group D) exhibited a sensitivity, specificity, positive predictive value, negative predictive value, positive likelihood ratio, and positive likelihood ratio of 0.442, 0.941, 0.826, 0.727, 7.512, 0.593, respectively, in predicting poor outcome.

### Interrelationship between overhydration status, being nutritionally at-risk, and sarcopenia

Figure 2 shows the interrelationship between overhydration, being nutritionally at-risk, and sarcopenia. Ten of 13 patients (76.9%) with the three comorbidities had a poor outcome. Patients with two of the three had a poor outcome in >50% cases, and patients with one of the three had a poor outcome in <50% cases. Fifty-six patients had none of the three factors; 10 (17.9%) of these had poor outcomes.

**Figure 2.**
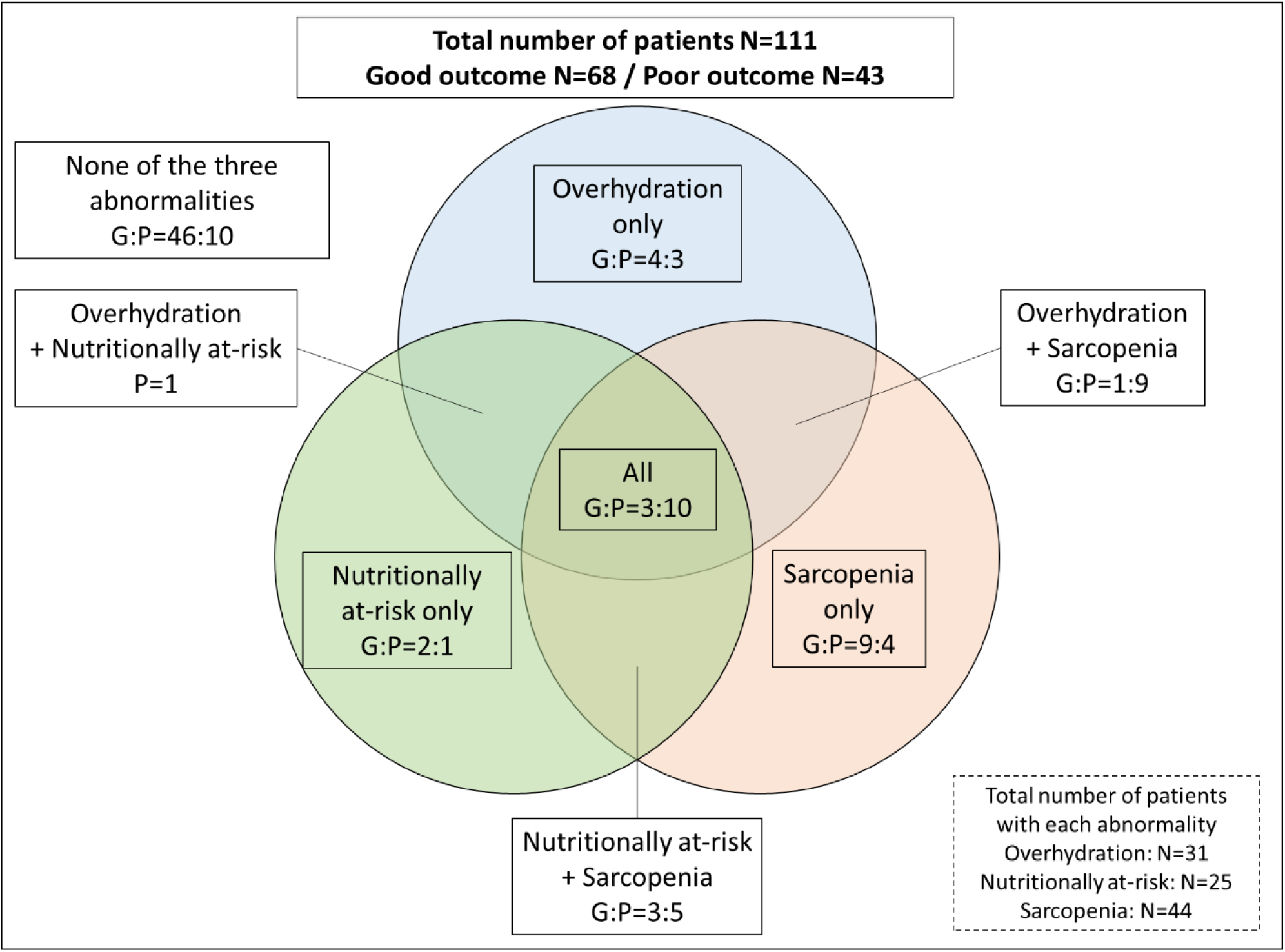
Interrelationship between overhydration, being nutritionally at-risk, and sarcopenia. Numbers indicate the number of patients. G, good outcome; P, poor outcome

### Multivariate analysis

Logistic regression analysis was performed with the good and poor outcome groups as dependent variables and age, sex, NIHSS score, NT–pro-BNP, overhydrated status, nutritionally at-risk, and sarcopenia as independent variables. The analysis revealed age (odds ratio [OR] 1.062, 95% confidence interval [CI] 1.010–1.117; P = 0.020), NIHSS score (OR 1.790, 95%CI 1.307–2.451; P < 0.001), and overhydration (OR 5.504, 95%CI 1.717–17.648; P = 0.001) as independent predictors of a poor outcome. The P value for this model was 0.219 with the Hosmer–Lemeshow test. In a sensitivity analysis, the model was analyzed with BMI added as an independent variable, albumin instead of nutritionally at-risk, and decreased grip strength and low muscle mass instead of sarcopenia, yielding similar results.

## Discussion

In the present study focusing on patients with AIS living independently before AIS onset, we evaluated the contribution of abnormal body water balance, especially overhydration, nutritional risk, and sarcopenia to poor outcomes at discharge. Although these three factors are interrelated,^12–14^ this study suggested the higher overlap among these factors, the higher the rate of poor outcome. In addition, the concomitant presence of overhydration and low muscle mass was highly specific for poor outcome after AIS. Our multivariate analysis revealed that overhydration, but not nutritionally at-risk or sarcopenia, was a predictor of poor outcome independent of age and stroke severity.

Human body is composed of four major components, namely fat, extracellular solids (ECS), extracellular fluid (ECF), and body cell mass (BCM), as illustrated in Figure 3. ^8,29^

**Figure 3.**
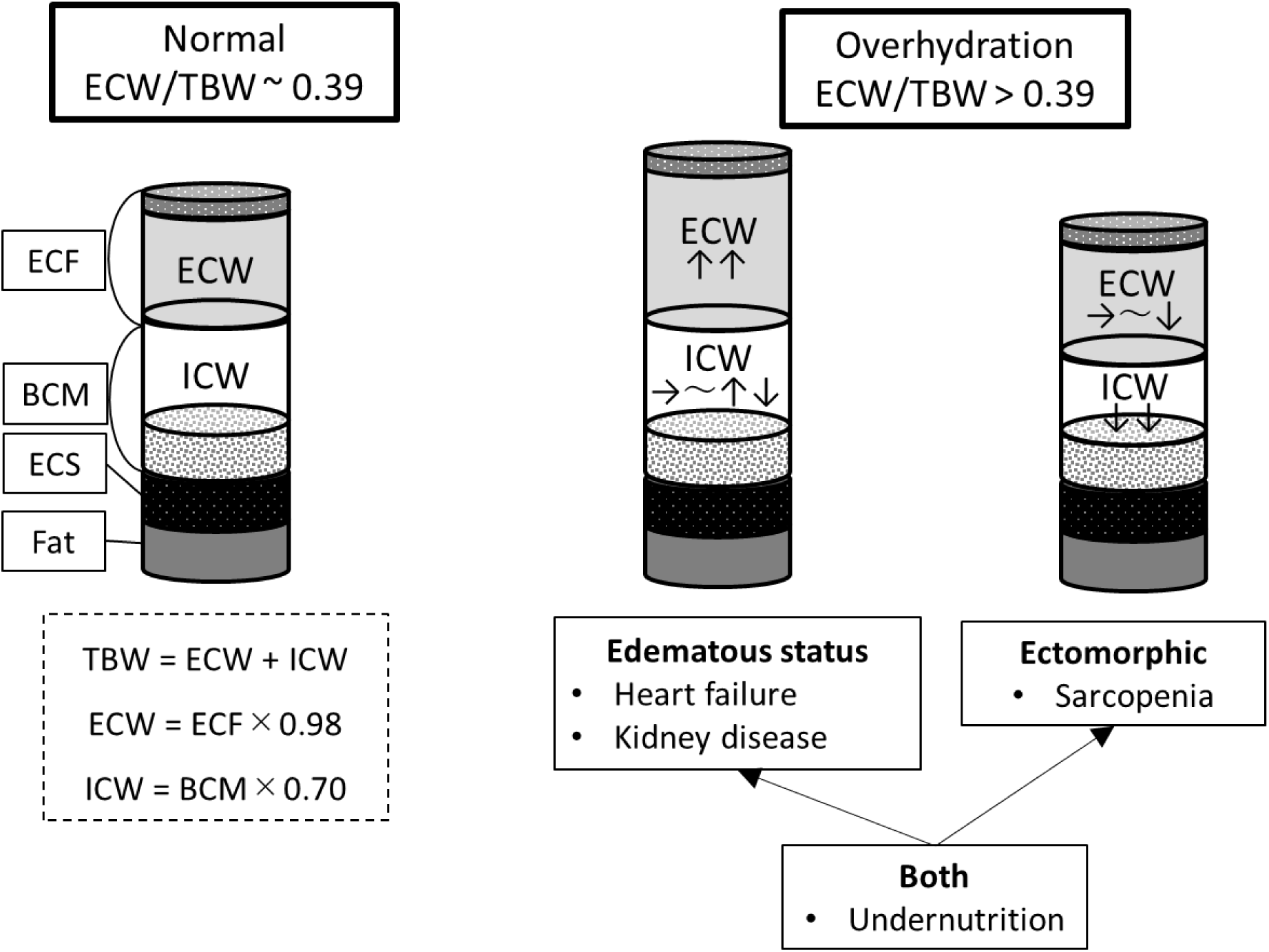
Body composition and conditions that cause overhydration BCM, body cell mass; ECF, extracellular fluid; ECS, extracellular solids; ECW, extracellular water; ICW, intracellular water; TBW, total body water

Among the four body components, fat consists of approximately 75% lipids with only a small amount of water.^30^ Unless the patient is morbidly obese or has undergone rapid weight change,^31,32^ the effect of fat water content on body water balance is minimal. ECS form a non-metabolizable component composed of organic and inorganic compounds such as bone matrix, collagen, and reticular and elastic fibers.^8,33^ ECS do not contribute to alterations in water balance as the ECS/TBW ratio is constant. ^8,33^ Conversely, there are two important factors related to body water balance: ECW and ICW. ECF represents fluid in circulating plasma and interstitium;^34^ 98% of the ECF is water mass, which is represented by ECW.^8^ On the other hand, BCM is primarily composed of water and protein; water mass accounts for approximately 70% of the BCM, which is represented by ICW.^8^ Cells are sensitive to changes in water content, and cellular swelling and shrinkage due to osmotic changes are associated with water movement to and from the ECW.^8,29^ This mechanism maintains the ICW/ECW ratio within a certain range in cases of water imbalance, and its failure leads to abnormalities in water balance.^8,29,35^ In other words, the ECW and ICW are body spaces allowing dynamic changes in the water balance.

Two pathogenic mechanism should be considered in the interpretation of the elevated ECW/TBW ratio observed. TBW is calculated as the sum of ECW and ICW; therefore, the increase in the ECW/TBW ratio can be ectomorphic, wherein the decrease in ICW is greater than the decrease in ECW, or edematous, wherein ECW increases while ICW remains unchanged or changes slightly (Figure 3).^29^ The extent to which ectomorphic and edematous states are involved varies across disorders. For example, the edematous state (i.e., increased ECW) plays a more significant role in increased ECW/TBW ratio in patients with acute heart failure^36^ and kidney disease.^22^ However, the involvement of the two pathogenic mechanism in AIS patients has not been determined. Skeletal muscle mass accounts for about half the mass of fat–free mass (ECF + BCM + ECS) in the human body.^8^ Approximately 79% of the weight of skeletal muscle is accounted for by water mass, which belongs to the ICW.^8^ Therefore, loss of muscle mass results in a decrease in ICW, and is related to diagnosis of sarcopenia.^3,4^ The prevalence of sarcopenia was higher in patients with a higher ECW/TBW ratio in the present study. This result indicated that sarcopenia accompanying AIS might have contributed to overhydration.

The GNRI score and muscle mass were lower in patients with a poor outcome, overhydration, and sarcopenia. Therefore, the poor outcome group appears to be not only at higher nutritional risk but also of being undernourished. Undernutrition here refers to protein–energy undernutrition (PEU), and edema appears in patients with PEU.^37^ In the present study, overhydration might have been influenced to some extent by edema due to undernutrition, in addition to the decreased ICW due to sarcopenia. In other words, the increase in the ECW/TBW ratio found in patients with a poor outcome after AIS could be related to both ectomorphism and edematous state. Our multivariate analysis indicated that the association of outcome with overhydration was stronger than its association with sarcopenia or nutritionally at-risk, which might be due to the fact that the ECW/TBW ratio reflects not only the loss of muscle mass due to sarcopenia but also the effects of undernutrition with edema. Indeed, the high positive predictive value of a high ECW/TBW ratio and a low muscle mass (Group D, Figure 1) for poor outcome suggests that both muscle mass and the ECW/TBW ratio might be useful in predicting patient outcomes after AIS in acute care hospital.

A study in rehabilitation units showed that most stroke patients suffer from poor nutritional status and loss of muscle mass during the acute phase of stroke.^27^ Therefore, nutritional management and early rehabilitation to maintain muscle mass are important.^27^ In the present study, patients with overhydration, nutritionally at-risk, and sarcopenia at the time of hospitalization had a higher rate of poor outcomes. This suggested that in addition to AIS treatments such as antithrombotic therapy, interventions targeting overhydration and undernutrition or nutritional risk and sarcopenia are important. Since a high intake of energy and protein, along with sufficient rehabilitation time, can effectively address sarcopenia related to AIS,^27,38^ these interventions may also help correct outcome in patients with AIS even in an acute care hospital.

Our findings should be interpreted with consideration of its limitations. First, this was a single-center study including a limited number of patients. Second, ECW and TBW, measured by BIA, slightly differ by race and ethnicity.^30,39^ In the present study, most of the patients were Japanese, with few Chinese patients, and the diagnostic criteria for sarcopenia used in Asia were considered in the analyses. Therefore, future studies should evaluate whether our findings are comparable in other ethnic groups or in the setting of different sarcopenia diagnostic criteria. Third, the diets and the intravenous fluids administered to the patients with AIS after admission were not evaluated. Evaluation of patients on specific days or hydration after hospitalization would provide further information in this regard.

## Conclusions

AIS patients with a poor outcome at discharge showed more overhydration, were nutritionally at-risk, and had sarcopenia. The higher the overlap of these three factors, the higher the rate of poor outcomes. Patients showing both overhydration and low muscle mass had particularly high rates of poor outcome. Overhydration, rather than sarcopenia or being nutritionally at-risk, was associated with poor outcome by multivariable analysis. Assessing body composition with BIA was useful in predicting outcomes after AIS in an acute care hospital.

## Data Availability

The data that support the findings of this study are available from the corresponding author upon reasonable request.

## Non-standard Abbreviations and Acronyms

AIS: acute ischemic stroke
BIA: bioelectrical impedance analysis
BCM: body cell mass
ECF: extracellular fluid
ECS: extracellular solids
ECW: extracellular water
GNRI: geriatric nutritional risk index
GLIM: Global Leadership Initiative on Malnutrition
IBW: ideal body weight
ICW: intracellular water
TBW: total body water

## Acknowledgments

The authors thank Yuki Yokota, Satoshi Hirose, and Naotoshi Natori for their assistance with the collection of individual data.

## Supplemental Material

Tables S1–S2

Figure S1

